# Household Air Pollution from Cooking Fuels and its Association with Under-Five Mortality in Bangladesh

**DOI:** 10.1101/2022.05.20.22275373

**Authors:** Md. Badsha Alam, Supria Acharjee, S. M. Ashique Mahmud, Jesmin Akter Tania, Md Mostaured Ali Khan, Md. Syful Islam, Md. Nuruzzaman Khan

## Abstract

**Background:** Solid fuel use was found to be associated with under-five mortality in low- and lower-middle income countries (LMICs). However, the current understanding of this association for Bangladesh is absent through around 80% of the total households in Bangladesh used solid fuel and the higher under-five mortality rate is a significant public health burden. We explored the associations of household cooking fuels used and the level of exposure to HAP through cooking fuels used with neonatal, infant, and under-five child mortality in Bangladesh.

**Methods:** We analysed 4,198 mother-child dyads data extracted from the 2017/18 Bangladesh Demographic and Health Survey data. Type of cooking fuels used (solid fuel, clean fuel) and level of exposure to HAP through cooking fuels used (unexposed, moderately exposed, highly exposed) were considered as exposure variables. Neonatal, infant and under-five mortality were considered as the outcome variables. Relationships between the exposure and outcome variable were explored by using the multilevel mixed-effect logistic regression model adjusting for possible confounders.

**Results:** Solid fuels were found to be used in nearly 80% of the total mothers analysed. A higher likelihood of mortality was found among neonates (aOR, 3.78; 95% CI, 1.14-12.51) and infants (aOR, 2.93; 95% CI, 1.60-6.15) of the women who used solid fuels as compared to the mothers who used clean fuel. The association was found strongest when we considered solid fuel used together with place of cooking. The likelihood of neonatal and infant mortality was found 4.33 (95% CI, 1.16-16.23) and 2.05 (95% CI, 1.18-7.23) times higher among mothers who were highly exposed to solid fuel used as compared to the mothers who were unexposed to solid fuel used.

**Conclusion:** Solid fuel used and its increased exposure increased the occurrence of neonatal and infant mortality. Prioritizing the use of clean fuel use and increasing awareness about the adverse effects of solid fuel use are important.

## Introduction

Household air pollution (HAP) is a significant public health concern in low- and lower-middle-income countries (LMICs) [1, 2]. It causes in several routes, however, the use of solid cooking fuels, e.g., peat, wood and coal, is the most common route. About 2.6 billion people worldwide use solid fuels for cooking, and a majority of them live in LMICs [3]. Consequently, of the 3.8 million premature deaths that occur worldwide every year because of HAP, almost all of them occur in LMICs [3]. HAP from solid fuel use was also found to be associated with several serve child morbidity, including childhood pneumonia, intrauterine growth restriction, preterm birth, and low birth weight [3-12]. There is also evidence that HAP use is linked with the rising rate of pregnancy complications, which further increase maternal hospitalization and caesarean section delivery [4-12].

Bangladesh has achieved significant progress in reducing under-five mortality during the Millennium Development Goals period of 2000 to 2015 [13]. However, the rate is still very high at 45 and 30 per 1000 live births for under-five mortality and neonatal mortality, respectively [13, 14]. This presents a challenge in achieving the Sustainable Development Goal 3 (health and wellbeing for all), in particular, its targets to reduce under-five (25 per 1000 live births) and neonatal (12 per 1000 live births) mortality rates by 2030 [13, 14]. It indicates the need to scaling up the current policies and programs to reduce under-five mortality by covering every possible area that causes under-five mortality.

In Bangladesh, as with other LMICs, solid fuels are the primary element of cooking or heating with around 80% prevalence. The rate is even higher for rural area (92%) [15]. Traditionally, women in LMICs including in Bangladesh are mainly involved in cooking activities and often they are accompanied by their young children at the time of cooking [16]. This makes young children highly vulnerable to HAP which could be linked to the current higher rate of under-five mortality in Bangladesh. This was found true in previous studies of Bangladesh conducted based on the data collected a decade ago from now along with other adverse consequences including low birth weight and pre-term birth [8, 17-21]. Moreover, available studies were also limited in several areas, including small geographic areas, small scale institutional-level data, and small sample sizes [8, 17-20, 22, 23].

Following this study period, a number of national-level policies and programs to increase awareness about the adverse effect of solid fuel use as well as to reduce the use of solid fuel have been taken in Bangladesh. As such, along with the country’s rapid socio-economic development, significant progress has been made in household cooking fuel using patterns, including rising dependency on electric and gas stoves [24]. Moreover, awareness of the adverse effects of solid fuel use also motivates people in changing their solid fuel using place, from inside to outside of the living room or corner of the yard. This could alternate or reduce the strength of the reported association between HAP and under-five mortality in Bangladesh. However, there is no recent evidence covering this issue. Moreover, evidence showing the association between the level of HAP exposure and under-five mortality has not been investigated in Bangladesh so far. However, this was even found as the strongest determinant of under-five mortality in other LMICs. Therefore, this association need to explore as a significant portion of the Bangladeshi population uses solid fuel in indoor places, such as in the living room or close to the living room, which further increases their HAP exposure level.

We conducted this study to fill these gaps. We explored the associations of solid fuel use and level of HAP from solid fuel use with under-five and neonatal mortality in Bangladesh by using the most recent nationally representative survey data. The findings will help the policymakers to know the current situation of the association between HAP and under-five mortality in Bangladesh. This will also be a learning point for other LMICs as like Bangladesh about whether the focus on HAP from solid fuel use could bring significant progress in reducing the country’s under-five mortality.

### Methods

### Study design and sample

Data for this study were extracted from the most recent Bangladesh Demographic and Health Survey (BDHS) conducted in 2017/18. The survey is conducted every three years as part of the Demographic and Health Survey (DHS) program of the USA. The National Institute of Population Research and Training (NIPORT), a government organization that works under the Ministry of Health and Family Welfare of Bangladesh, conducted this survey. International development partners including UNFPA, and USAID provide other supports including financial and technical support. The survey selected a list of nationally representative sample of households from where the eligible women (reproductive-aged married women who are a usual residents of the selected household or passed their most recent night at the selected household) were included. The households were selected by using two-stage stratified random sampling methods. At the first stage of sampling, 675 Primary Sampling Units (PSUs) were selected covering every administrative division of Bangladesh and rural and urban areas. The PSU was selected from a list of 293,579 PSUs of Bangladesh which was generated by the Bangladesh Bureau of Statistics as part of the 2011 Bangladesh National Population Census. A total of 672 PSUs was retained finally after excluding 3 PSUs because of extreme flooding. A fixed number of 30 households was selected at the second stage from each selected PSU. Finally, the survey was conducted in 19,457 households with an over 96% inclusion rate. There were 20,376 women in these selected households, of them, 20,127 women were interviewed with a response rate of 98.8%. A detailed description of the sampling procedure is available elsewhere [25].

### Study sample

We analysed 4,189 mother-child dyads data. The sample was extracted from the original sample based on the following inclusion criteria: (i) mothers had at least one birth within five years of the survey date, (ii) reported survival status of their children, and (iii) reported type of cooking fuels they used and the place where they cooked.

### Outcome variables

We considered three outcomes’ variables: neonatal mortality (death occurred within 1 month of live birth), infant mortality (deaths occurred within 12 months of live birth) and under-five mortality (death occurred within 60 months of live births). The BDHS recorded these mortality data by asking women whether she had any live birth within five years of the survey date and survival status of the respective child. In the occurrence of more than one life birth within five years, survival status data were collected for every child. These data were then recategorized by following the relevant guidelines of the WHO to generate outcomes variables.

### Exposure variables

Two exposure variables were considered: (i) type of cooking fuels use (solid fuel, clean fuel) and (ii) level of exposure to HAP through cooking fuels (unexposed, moderately exposed, highly exposed). The BDHS recorded the type of fuel that used in the respondents’ households for cooking through asking *“What type of fuel does your households mainly used for cooking?”*. A list of fuel was provided to give the response. In case, the fuel used by the respondents was not in the list, they were allowed to write the name of the fuel. We reclassified these responses as solid fuel used (if the respondents recorded coal, lignite, charcoal, wood, straw, shrubs, grass, agricultural crop and animal dung) and clean fuel used (electricity, liquid petroleum gas, natural gas and biogas) to generate the first exposure variable. Respondents were also asked about place of cooking in their households through asking *“Is the cooking usually done in the house, in a separate building, or outdoors?”*. Responses recorded for this question was considered along with the type of cooking fuel respondents used to generate the second exposure variable. The respondents were considered as *unexposed*, if respondents recorded clean fuels use in their households for cooking purpose; *moderately exposed*, if respondents recorded solid fuels use in their households for cooking purpose, however, the cooking conducts in a separate building or outdoors; and *highly exposed*, if respondents recorded solid fuels use in their households for cooking purpose, however, the cooking conducts inside their houses. We generate this variable by following previous studies conducted in LMICs [8, 17-19, 21, 26-28].

### Confounding adjustment

Individual-, household-, and community level factors were considered as confounding factors. We first generate a list of confounding variables by reviewing the relevant studies conducted in LMICs, particularly Bangladesh [8, 17-20, 22, 23, 29]. The availability of the data of the selected variables in the survey we analysed were then checked. The variables whose data were found to be available in the survey were then analysed with the forward regression analysis. The variables that were found significant, as presented in Table 3, were considered as confounding factors.

### Statistical analysis

Descriptive statistics was used to describe the characteristics of the respondents. Separate multilevel mixed-effect logistic regression model was used to assess the associations of type of cooking fuels use and exposure level of HAP through using solid fuel with neonatal, infant and under-five mortality. The reason for using this model was nesting structure of the BDHS data where the previous studies found multilevel modelling produce comparatively better findings [30]. Survey weight was also considered in the model. We runed both unadjusted and adjusted models. In the unadjusted model, only the particular exposure and outcome variable was considered. The confounding variables considered were added with the particular exposure and outcome variable in the adjusted model. Results were recorded as Odd Ratio (OR) with its 95% Confidence Interval (95% CI). We performed all descriptive statistics using the Stata software version 15.1 (Stata corporation, college station, Texas, USA).

## Results

Table 1 presents background characteristics of the mothers and under-five children. The mean (±SD) age of the mothers was 26.80 (±5.08) years and mean year of education was 5.79 (±3.68) years. The mean age of the children we analysed was 2.05 ((±1.44) years. Nearly 47 % of the total children analysed were girls.

**Table 1:**
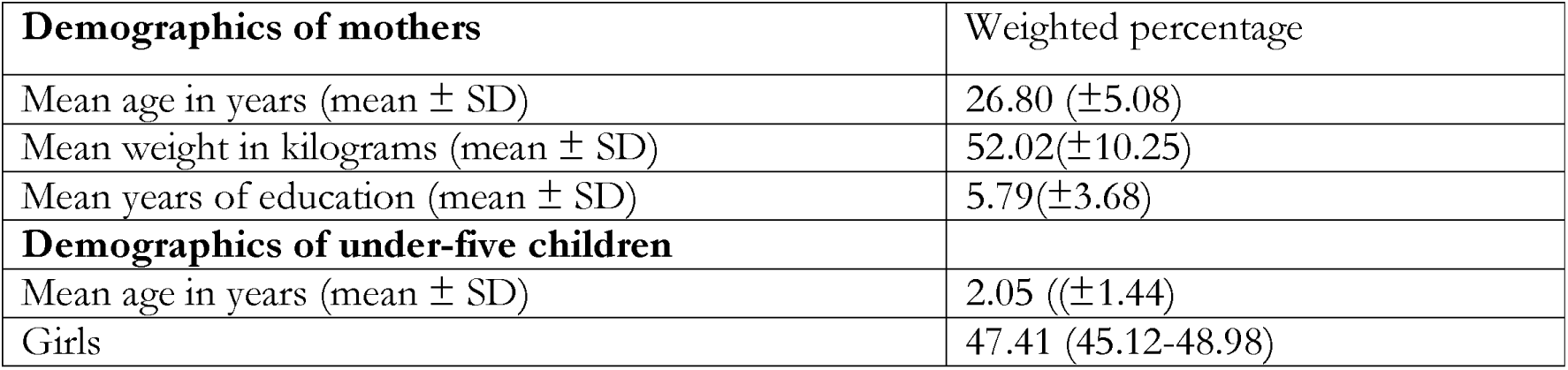
Background characteristics of the respondents (N=4,189)

The distribution of the type of cooking fuel used in the household and the place of household’s cooking are presented in Table 2. We found wood as major type of cooking fuel in Bangladesh with around 45% of the total use following agricultural crop (27%) and liquid petroleum gas/natural gas (19.11%). Nearly 3% of the total respondents analysed reported indoor cooking place. When we considered fuel using pattern together, we found solid fuels were used around 80% of the respondents’ households. Around 79% of the total respondents’ households were found to be moderately exposed to HAP through using solid fuel used and 1.21% of the total respondents’ households were found highly exposed to HAP through solid fuel used.

**Table 2:** Distribution of exposures and outcomes variables, BDHS, 2017/18 (N=4,189)

|  |  |
| --- | --- |
| **Types of cooking fuels** | Weighted percentage |
| Electricity | 0.37 |
| Liquid petroleum gas + natural gas | 19.11 |
| Charcoal | 0.11 |
| Wood | 44.79 |
| Agricultural crop | 27.12 |
| Coal, lignite + straw/shrubs/grass + others | 8.44 |
| **Cooking place** |  |
| Indoor | 2.77 |
| Outdoors | 97.23 |
| **Categorization of exposure variables** |  |
| **Type of cooking fuels used** |  |
| Solid fuel use | 80.48 |
| Clean fuel use | 19.52 |
| **Level of exposure to HAP through cooking fuels used** |  |
| Unexposed | 19.52 |
| Moderately exposed | 79.27 |
| Highly exposed | 1.21 |
| **Outcome variables** |  |
| Neonatal mortality per 1000 live births | 25.1 |
| Infant mortality per 1000 live births | 34.5 |
| Under-five mortality per 1000 live births | 37.8 |

The current distribution of the neonatal mortality, infant mortality and under-five mortality in Bangladesh are also presented in Table 2. We reported 38 under-five deaths per 1000 live births following infant mortality of 34 per 1000 live births and neonatal mortality of 25 per 1000 live births.

We also explored weighted percentage of neonatal, infant and under-five mortality across confounding variables considered in the analysis. Relevant results are presented in Table 3. A higher rates of neonatal, infant and under-five mortality was found among the children of the mothers who were aged 25-35 years, primary or secondary educated, moderately exposed to mass media and poor. Each form of mortality was also found higher among male children. A higher proportion of the neonatal, infant and under-five mortality was also found among the children of the rural mothers or mothers of the Rangpur and Dhaka division.

**Table 3:** Weighted percentage of neonatal, infant and under-five mortality by confounding variables, BDHS, 2017/18.

| **Maternal age** | **Neonatal mortality per 1000** | **Infant Mortality per 1000** | **Under-five mortality per 1000** |
| --- | --- | --- | --- |
| **Mothers' ages** |  |  |  |
| ≤ 24 years | 33.91 | 32.39 | 32.52 |
| 25–35 years | 61.72 | 64.42 | 64.44 |
| > 35 years | 4.37 | 3.18 | 3.04 |
| **Mother's education** |  |  |  |
| Illiterate | 14.11 | 12.35 | 14.06 |
| Primary | 36.48 | 38.05 | 37.22 |
| Secondary | 42.31 | 42.41 | 41.24 |
| Higher | 7.10 | 7.18 | 7.48 |
| **Child's gender** |  |  |  |
| Male | 57.34 | 56.88 | 54.61 |
| Female | 42.66 | 43.12 | 45.39 |
| **Birth interval** |  |  |  |
| ≤24 months | 18.83 | 15.31 | 15.05 |
| >24 months | 81.17 | 84.68 | 84.95 |
| **Exposure to mass media** |  |  |  |
| Not exposed | 37.53 | 38.58 | 41.60 |
| Moderately exposed | 55.43 | 55.16 | 52.69 |
| Highly exposed | 7.04 | 6.26 | 5.71 |
| **Household wealth quintiles** |  |  |  |
| Poor | 53.77 | 54.42 | 55.50 |
| Middle | 18.81 | 16.46 | 15.88 |
| Richer | 27.41 | 29.12 | 28.62 |
| **Urbanicity** |  |  |  |
| Urban | 29.96 | 28.83 | 27.30 |
| Rural | 70.04 | 71.17 | 72.70 |
| **Place of residence** |  |  |  |
| Barishal | 7.78 | 6.86 | 7.04 |
| Chattogram | 16.23 | 14.48 | 16.01 |
| Dhaka | 21.07 | 23.39 | 22.83 |
| Khulna | 10.80 | 9.44 | 8.61 |
| Mymensingh | 3.71 | 4.95 | 4.51 |
| Rajshahi | 11.35 | 11.99 | 13.06 |
| Rangpur | 20.36 | 15.42 | 15.09 |
| Sylhet | 9.00 | 13.47 | 12.85 |

**Note:** All estimated presented in this table are column percentage

The unadjusted and adjusted associations between the exposure and outcome variable are presented in Table 4. The likelihoods of neonatal mortality (adjusted OR (aOR) 3.78; 95% CI, 1.14–12.51) and infant mortality (aOR, 2.93, 95% CI 1.60–6.15) were found higher among mothers reported use of solid fuels as compared to the mothers reported use of clean fuel. The likelihood of neonatal mortality was even found more strongest when we considered solid fuel used together with place of cooking. We found 4.33 times (95% CI, 1.16-16.23) higher likelihood of neonatal mortality among neonates of the mothers classified as highly exposed to HAP through solid fuel used as compared to the mothers classified as unexposed to HAP through solid fuel used. However, for infant mortality, the odds was 2.05 times (95% CI, 1.18-7.23) higher among infants of the mothers classified as highly exposed to HAP through solid fuel used as compared to the mothers classified as unexposed to HAP through solid fuel used. We did not find any significant association of under-five mortality with exposure variables considered both in the adjusted and unadjusted models though the relevant odds were higher.

**Table 4:** Results of the multilevel mixed-effect logistic regression model in assessing the association of household cooking fuels used and level of exposure to HAP through cooking fuels used with the neonatal, infant, and under-five child mortality, Bangladesh, 2017/18.

| Exposure | Neonatal mortality<br>OR (95% CI) | Infant mortality<br>OR (95% CI) | Under-five<br>mortality<br>OR (95% CI) |
| --- | --- | --- | --- |
| **Unadjusted association** |  |  |  |
| **Type of cooking fuels used** |  |  |  |
| Clean fuel (ref) | 1.00 | 1.00 | 1.00 |
| Solid fuel | 3.44 (1.17-10.13)** | 2.39 (1.03-6.84)** | 2.08 (0.73-5.93) |
| **Level of exposure to HAP through cooking fuels used** |  |  |  |
| Unexposed (ref) | 1.00 | 1.00 | 1.00 |
| Moderately exposed | 1.09 (0.63-1.88) | 1.02 (0.64-1.62) | 1.12 (0.71-1.77) |
| Highly exposed | 3.76 (1.15-12.33)** | 2.45 (1.43-7.66)** | 2.30 (0.73-7.27) |
| **Adjusted association++** |  |  |  |
| **Type of cooking fuels used** |  |  |  |
| Clean fuel | 1.00 | 1.00 | 1.00 |
| Solid fuel | 3.78 (1.14-12.51)** | 2.93 (1.60-6.15)** | 1.74 (0.55-5.46) |
| **Level of exposure to HAP through cooking fuels used** |  |  |  |
| Unexposed | 1.00 | 1.00 | 1.00 |
| Moderate | 1.05 (0.58-1.87) | 0.98 (0.61-1.58) | 1.10 (0.69-1.75) |
| High | 4.33 (1.16-16.23)** | 2.05 (1.18-7.23)** | 1.99 (0.57-6.99) |

**Notes:** \*\*p<0.05, ++Models are adjusted for mothers' age, mothers' education, child gender, birth interval, exposure to mass media, household wealth quintile, urbanacity and place of residence.

## Discussion

In this nationally representative study by analysing the most recent 2017/18 BDHS data, we examined the associations of household cooking fuels used and the level of exposure to HAP through cooking fuels used with the neonatal, infant, and under-five child mortality in Bangladesh. We found solid fuels were used by nearly 80% of the mothers’ households and nearly 3% used indoor places for cooking. The rate of under-five, infant and neonatal mortality was found around 38, 34, and 25 per 1000 live births, respectively. We found a higher probability of neonatal and infant mortality in mothers who used solid fuels in their households and were classified as highly exposed to HAP through solid fuel used, compared to their counterparts who reported using clean fuels in their households and not were exposed to HAP through solid fuel used, respectively. On the other hand, associations between under-five mortality and fuel using type and place were not found significant. These findings are robust in response to the research data analysed, the type of confounding factors considered, and the statistical method used. Therefore, the findings are expected to help the policymakers to create evidence-based policies and programs to achieve SDGs targets of reducing under-five and neonatal deaths in Bangladesh by 2030.

As reported in this study, infant and under-five mortality rates are higher in rural areas and among poor mothers. These are identical to the findings of other studies conducted in Bangladesh [8, 14, 31]. In terms of the place of residence, neonatal, infant and under-five mortality rates were found higher among children of rural mothers or mothers residing in the Dhaka and Rangpur division. They contradict with the available studies’ findings conducted by using the 2014 BDHS data while the prevalence of child mortality was found higher in the Sylhet division [8, 14].

Previous studies in Bangladesh reported HAP because of using solid fuels was associated with an increased risk of neonatal and infant mortality [21, 26], which coincides exactly with this updated analysis. Studies conducted in other LMICs like Myanmar and India also reported a similar association [27, 32]. However, the current association is found stronger than the previous evidence from Bangladesh as this study used the most updated and rigorous statistical methods with a potential list of confounders [8, 23]. For the first time in Bangladesh, we also found likelihoods of neonatal and infant mortality increased with the increasing level of exposure to HAP through solid fuel used. These associations were consistent with the recently reported association for Myanmar [27].

Respiratory systems of the neonatal and infants are comparatively weak and they comparatively breathe a higher volume of air [16]. Even if solid fuels are being used indoors it drastically increases the airborne toxic pollutants’ concentration in the household and ambient air [8, 21]. These increase occurrence of respiratory diseases, including pneumonia, chronic bronchitis, and asthma [3, 33]. The occurrence of such morbidities at a very early stage then led to an increased risk of mortality. This mechanism can also explain our reported findings of insignificant associations between under-five mortality and solid fuel use. The respiratory system of the under-five aged children is comparatively strongest than that of neonates and infants [34]. Moreover, with increasing age, mothers may leave their under-five children to other family members before entering the cooking place. Such a comparatively strong respiratory system and lower exposure to HAP through solid fuel use can then lead to the insignificant association between solid fuel use and under-five mortality.

Our results suggest that the likelihood of neonatal and infant mortality increased with the increased exposure to HAP through solid fuel used. The effect size was even higher than the effects of solid fuel used on neonatal and infant mortality. This is possibly due to the fact that the effects of fumes from solid fuel are substantially higher when they are inhaled during indoor cooking [8]. Moreover, poor people are usually considered the indoor cooking places because of their poverty they could not make their houses with enough space to make a separate room for cooking [20]. Additionally, the indoor cooking places usually does not have adequate fumes ventilation systems [8]. Besides these, lower breastfeeding, immunization, child malnutrition, and inappropriate or inadequate food taking behaviour are also found higher among poor people in Bangladesh as well as other LMICs. Therefore, their pooled effects could be responsible for the rising occurrence of neonatal or infant mortality with the increased exposure to HAP through solid fuel used.

This study has several strengths and some limitations. First, we analysed large-scale nationally representative research data that is desirable for policy and programs making at the national level. Advanced statistical modelling was used in this study, along with a precious list of confounding factors. Therefore, the current research results are the most accurate than the available findings. The major limitation of this study is analysis of the cross-sectional data; therefore, the findings are correlational only rather than casual. All data were self-reported, therefore, recall biased may raise. However, any of such bias is likely to be random. Given the nature of the association we explored, the reasons for child deaths to be occurred should be considered important for adjustment. However, the survey did not record such information, as such, we could not adjust this in the model. Kitchen ventilation system should also be considered important for adjustment for the association we measured. However, this data was also not available in the survey we analysed.

## Conclusion

Solid fuel use and highly exposed to HAP through solid fuel used were found to be associated with neonatal and infant mortality, but not with under-five mortality. Considering current 80% of the total Bangladeshi households use solid fuel and cooking fuel is a preventable risk factor that can be modified through developments in housing design, health-care policies, infrastructure, and behavioral interventions, this study findings indicates needs for serious attention from the policy makers to make policies and programs to increase awareness about the adverse effects of solid fuel used. Special focus needs to be given for the mother having under-five aged children. Behavioral interventions, such as promoting the use of improved cooking stoves, increased natural household ventilation, and not carrying babies while cooking, are likely to play an important role in reducing neonatal and infant mortality in Bangladesh.

## Declaration

This study has not been published or presented anywhere before.

I confirm that the manuscript has been submitted solely to this journal and is not published, in-press, or submitted elsewhere.

I can confirm that all the research meets the ethical guidelines, including adherence to the legal requirements of the study country.

## Ethics

This study analysed secondary data publicly available. Ethical approval for this survey was provided by the Bangladesh Medical research counsel and Demographic and Health Survey Program of the USA. No additional ethical approval is required to conduct this study.

## Abbreviations

LMICs: Low- and Lower-Middle-Income Countries
SDGs: Sustainable Development Goals
BDHS: Bangladesh Demographic and Health Survey
aOR: Adjusted Odds Ratio, 95%
CI: 95% Confidence Interval

## Consent for publication

Not applicable

## Availability of data and materials

The Demography and Health Survey (DHS) program of the USA is the custodian of 2017 BDHS data. It is freely available for the user upon submission reasonable request to the DHS.

## Authors’ contribution

Khan MN, Islam MS, and Khan MM designed this study. Alam MB and Acharjee S analyzed the data along. Alam MB, Mahmud A, and Tania JA write the first draft of this manuscript. Khan MN, Islam MS, and Khan MM critically revised this manuscript. All authors approved this submitted version of the manuscript.

## Acknowledgement

We acknowledge DHS program of the USA, custodian of the data used in this study, for approving to use their data.

